# IL-18 and Lower Risk for Lung Cancer: Triangulated Evidence from Germline Predictions, Pre-Diagnostic Measurements, and Tumor Expression

**DOI:** 10.1101/2021.03.26.21254400

**Authors:** Karl Smith-Byrne, Yan Chen, Linda Kachuri, Pooja M. Kapoor, Florence Guida, Hana Zahed, Karine Alcala, Christopher Amos, Joshua Atkins, Barbara Bodinier, Aida Ferreiro-Iglesia, Rayjean J. Hung, Mikael Johansson, Woon-Puay Koh, Cecilie Kyrø, Arnulf Langhammer, Maik Pietzner, Núria Sala, Torkjel M. Sandanger, Ruth C. Travis, Kostas Tsilidis, Jian-Min Yuan, Hilary Robbins, Paul Brennan, James McKay, Mattias Johansson, Anders Mälarstig

## Abstract

Lung cancer remains the most common cause of cancer death globally. Dysregulation of immune response and inflammatory signaling is known to play an important role in lung tumorigenesis, but the causal drivers of this process have yet to be elucidated. To identify circulating inflammatory and immune-related proteins that influence risk for lung cancer we related genetically predicted plasma levels for 85 inflammation and immune proteins with susceptibility to lung cancer. Mendelian randomization (MR) analyses in 29,266 cases and 56,450 controls identified a candidate causal marker, IL-18, which conferred lower risk of lung cancer (OR per standard deviation increase: 0.85 [95% CI: 0.79-0.92]), in particular for adenocarcinoma (OR: 0.80 [95% CI: 0.72-0.89]). We subsequently validated this association using polygenic IL-18 predictions in the UK Biobank (HR highest vs. lowest quartile: 0.83 [95% CI: 0.72-0.95]) and using pre-diagnostic blood concentrations of IL-18 in 732 cases and 732 controls after controlling for the inhibitory role of IL-18BP (OR highest vs. lowest quartile: 0. 63 [95% CI: 0. 41-0.91]). Genetic colocalization suggested that IL-18 may act on lung cancer risk locally via lung tissue expression, and joint MR and tumor microenvironment analyses highlight CD8 T cells and NK cells as potential mediators. In addition to risk, IL-18 expression in adenocarcinoma tumor tissue was found to be associated with all-cause mortality in 480 TCGA samples after controlling for IL-18BP (HR per SD: 0.87 [95% CI: 0.78, 0.98]), which is in line with recent studies showing anti-tumor effects of IL-18. Our comprehensive genomic triangulation study thus highlights the potential for IL-18 as an aetiological biomarker and targetable for immune-oncology therapies.

## Introduction

Lung cancer is the third most highly incident cancer and the leading cause of cancer death globally^1^. While tobacco exposure accounts for most lung cancer diagnoses^2^, a majority of lung cancers in western countries are now diagnosed among former and never smokers^3^. Identifying novel aetiological and modifiable risk factors that aid the prevention of lung cancer is therefore important and should be prioritised.

The role of inflammation in lung cancer aetiology, such as IL-6^4,5^, IL-8^5^ and the IL-1 beta pathway^6^, presents an attractive potential target for chemoprevention due to their role in tissue remodelling and homeostasis^7^, cell proliferation^8^, and angiogenesis^9^. However, despite consistent observational evidence for the role of specific inflammation and immune-related proteins^4,5,10^, the confounding role of smoking and other biases not well-remedied through traditional epidemiology have precluded the identification of an aetiological role for inflammation in lung cancer aetiology.

Modern molecular epidemiology may overcome such concerns by integrating evidence from multiple, independent, lines of evidence drawn from study designs with differing sources of bias, such as Mendelian Randomisation (MR), well-designed prospective studies, and patient tumor characteristics – an approach commonly referred to as *triangulation*^11,12^. MR leverages common genetic variation to instrument the association of an exposure (such as inflammatory protein levels) with an outcome (lung cancer). The random segregation of alleles at conception requires that genetic variation be assigned prior to disease onset and substantially reduces the influence of biases that plague classical epidemiology. Nonetheless, MR remains sensitive to a number of assumptions, such as horizontal pleiotropy where a genetic instrument may associate with the outcome via both an exposure of interest and an alternative exposure like smoking. In the case of proteins, such bias may be alleviated by using genetic variation located adjacent to its coding gene that influence circulating concentrations by affecting transcription or translation^13^. Where analyses also preclude confounding by linkage disequilibrium (LD), MR represents a strong complementary source of evidence for the discovery of novel aetiological factors. For instance, a protein associated with lung cancer risk using MR and a well-designed prospective epidemiological study, with additional support from tumour characteristics (e.g. gene expression, gene-specific mutation etc), would provide compelling evidence for a role in aetiology not likely explained by a single source of confounding, such as tobacco exposure.

We aimed to identify novel inflammation and immune-related proteins in lung cancer aetiology. Following an initial MR-based discovery analysis, we sought to triangulate promising protein-risk signals using polygenic protein predictions in an independent large prospective cohort, direct pre-diagnostic measurements from a case-control study, and molecular and immune cell characteristics from patient tumor samples.

## Methods

### Overall study design

An overview of the study design is shown in Figure 1. Our objective was to identify aetiological inflammation and immune-related proteins for lung cancer risk using complementary sources of data. Based on genetic instruments identified in a protein GWAS of 30,913 individuals, we initially conducted an exploratory two-sample MR using a large GWAS to identify candidate proteins. Proteins with support from MR after sensitivity and colocalization analyses, were evaluated in relation to the tumor immune microenvironment to assess potential pathways using both MR and estimated tumor immune characteristics among lung cancer patients. Subsequently, we sought additional support for the importance of promising proteins with lung cancer using polygenic predicted protein concentrations in the UK Biobank, direct measurements in pre-diagnostic blood from a case-control study nested in six large population cohorts, and tumor gene expression for promising proteins with patient prognosis in the Cancer Genome Atlas^14^ (TCGA). STROBE-MR^15^ and STROBE^16^ reporting guidelines were followed.

**Figure 1.**
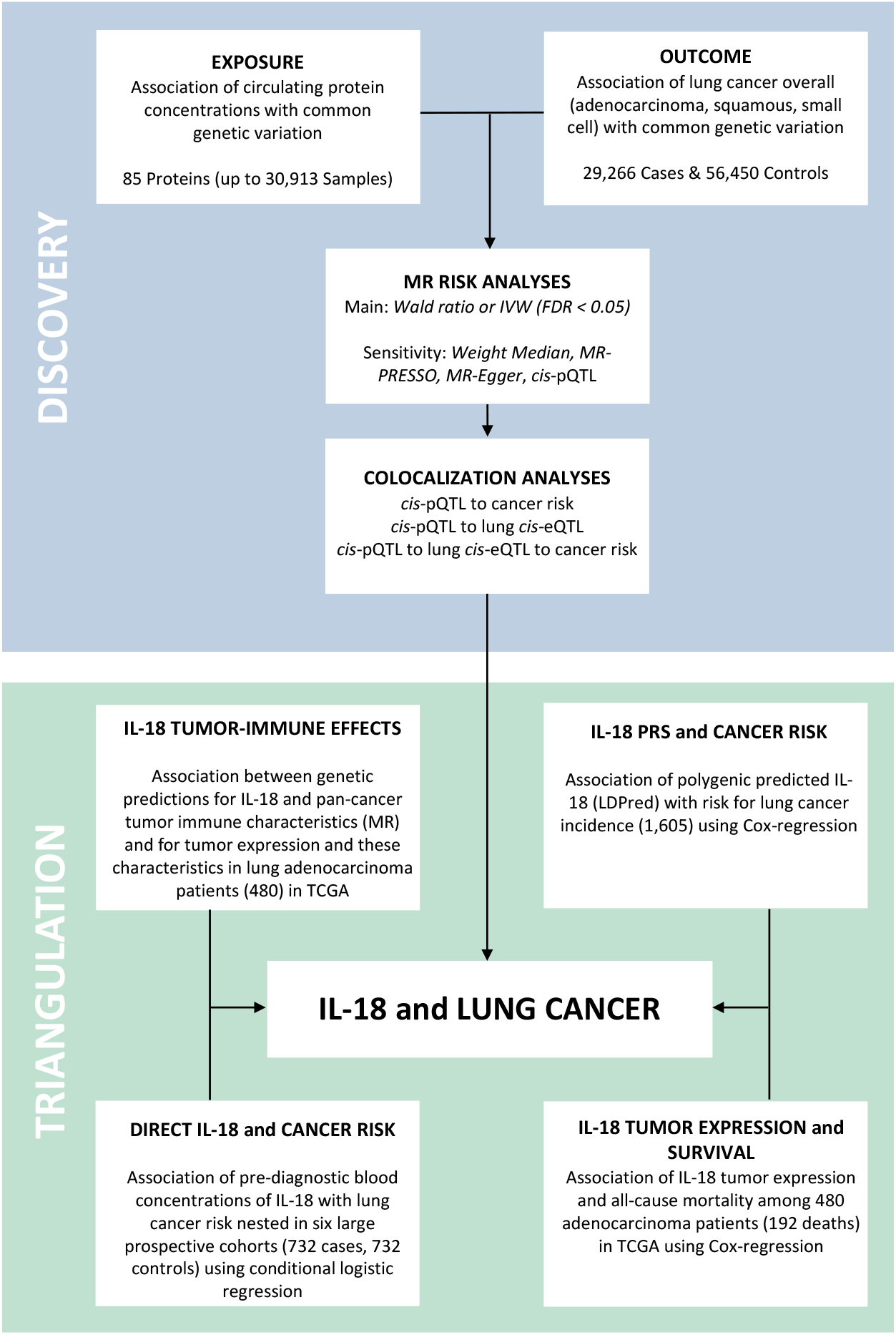
Overall study design. The objective of this study was to identify support for inflammation and immune-related proteins involved in the aetiology of lung cancer using a Mendelian randomization (MR) discovery subsequently triangulated using complementary evidence from independent source of data. In the MR discovery we find strong statistical support for the association of IL-18 with risk for lung cancer. Additionally, colocalization analyses may suggest that the association for IL-18 and lung cancer my act via expression in lung tissue. Following the MR discovery, we sought support for IL-18 and lung cancer risk using polygenic genetic IL-18 predictions in the UK Biobank, pre-diagnostic blood measurement in a large prospective nested case-control, and patient tumor characteristics and all-cause mortality. In these data we found broad support for IL-18 and lung cancer risk and patient mortality, albeit only after taking into account the inhibitory effect of IL-18BP for blood and tumor measurements. Additional MR and individual-level analyses for the association of IL-18 with tumor immune characteristics highlight CD8 T cells and NK cells as potential intermediate phenotypes.

## Identification of promising lung cancer proteins

### Mendelian randomization Risk Analyses

#### Study populations and data sources

Summary statistics for the association of SNPs with protein concentrations were obtained from a recent protein GWAS in 30,931 subjects of European descent from 14 studies within the SCALLOP consortium of the Cardiovascular-I panel from Olink Proteomics^17^. SNP-lung cancer risk associations were extracted from a recent meta-analysis of the International Lung Cancer Consortium (ILCCO) combining results from a large-scale lung cancer GWAS with 29,266 cases and 56,450 controls of European descent^18^. All studies received ethical approval from their respective review committees/boards and all participants provided written informed consent.

#### Main Statistical analysis

Of the 90 proteins assayed in 30,931 individuals with genome-wide genotyping data, 85 circulating proteins had SNPs associated at genome-wide significance (*P*<5×10^−8^), of which 74 had *cis*-protein quantitative trait (pQTL)s located within 1mb of the protein-coding gene. Only SNPs that were independent (linkage disequilibrium, *r*^*2*^ > 0.01) from other SNPs for a given protein, and with no significant heterogeneity of SNP effects across the 14 studies contributing to GWAS meta-analysis were included in further analyses. After data harmonization, 85 proteins with 366 unique SNPs, including 129 *cis-*pQTL, were included in analyses. There was no overlap of participants between the protein and lung cancer GWAS samples.

In MR analysis to describe the association between proteins and risk for lung cancer, for proteins with one available robust pQTL SNP available we used Wald estimates (β_cancer_/β_protein_), and for proteins with multiple pQTL SNPs per-SNP Wald estimates were combined using the inverse-variance weighted approach (IVW). A false discovery rate (FDR) of 5% was applied to define a p-value threshold corrected for multiple testing to identify promising risk proteins for downstream analyses.

We performed quantitative and qualitative sensitivity analyses to evaluate potential violations of MR assumptions. For proteins with FDR significant Wald/IVW estimates, we ran weighted-median^19^, MR-PRESSO^20^, and MR-Egger^21^ sensitivity analyses that provide estimates more robust to bias from horizontal pleiotropy and can quantify net directional pleiotropy (using the MR-Egger intercept). Heterogeneity of the SNP estimates—an indication of horizontal pleiotropy—was evaluated using Cochran’s and Rücker’s Q^22^. Subsequently, we consulted all phenotypes in the OpenGWAS^23^ catalogue to identify potential horizontal pleiotropy for pQTL instruments used in main MR analyses. As an additional assessment of pleiotropy, we conducted a proteome-wide association analysis across 4,979 proteins (multiple testing correction, p<1e-5) for the *cis* and *trans* pQTL used to estimate risk associations for proteins otherwise passing sensitivity analyses. These analyses were in 8,350 participants using linear regression adjusted for age, sex, center, and the first ten genetic principal components (data not shown) within the Fenland study^24^.

#### Cis-pQTL MR and Colocalization analyses

For proteins passing these MR sensitivity analyses and with available *cis*-pQTL (i.e. SNPs in or within 1Mb of the protein-coding gene), a separate MR analysis was conducted using the Wald ratio to estimate risk. Further, we performed colocalization^25,26^ analyses to evaluate confounding due to linkage disequilibrium using a 75kb region up- and downstream of a given *cis*-pQTL. Bidirectional MR was used to estimate potential lung cancer effects on protein concentrations as a sensitivity analysis to assess the correct orientation of MR estimates.

#### Triangulating evidence for promising lung cancer proteins Protein-Tumor Immune Analyses

To evaluate the influence on the tumor immune landscape for the promising lung cancer proteins identified through MR, we conducted analyses using both MR and individual level patient tumor characteristics based on patients within TCGA. To conduct MR analyses, summary statistics for each of the *cis* and *trans* pQTL used in the main risk analyses were extracted for tumor immune characteristics from a recent pan-cancer GWAS that included ∼9,000 cancer patients from 30 cancer subtypes from TCGA^27^. All MR methods applied were as described above. Analyses in individual-level data for the association of lung tissue gene expression for promising proteins with tumor immune characteristics were conducted on data obtained using TCGA project 2731 via dbGAP. 480 adenocarcinoma and 420 squamous cell carcinoma patient samples remained after filtering on annotations and removing duplicated samples (Supplementary Methods). Linear regression adjusted for age at diagnosis, sex, smoking status, and tumor stage were used to estimate the association of gene expression with tumor immune characteristics.

#### Protein PRS and Lung Cancer Risk

We used polygenic predictions (PRS) calculated using the LDPred algorithm^28^ to estimate the association for promising proteins with lung cancer risk using individual-level data from the UK Biobank. PRS typically explain a greater proportion of the variance for protein concentrations than GWAS significant hits. Therefore, they may be better powered to discover associations with the caveat that they cannot be understood as causal estimates, but rather as a complementary analysis to MR in independent individual-level samples. The UK Biobank is a large ongoing prospective cohort, from 2006, with more than 500,000 participants described in Bycroft et al.^29^ and accessed under project number 15825 (Supplementary Methods). Cox regression was used to estimate the association of genetically predicted protein concentrations with lung cancer risk in UK Biobank. All models were adjusted for age, genotyping array, and the first five principal components of ancestry, and stratified by sex. Sensitivity analyses were also stratified by smoking status.

#### Pre-diagnostic Protein Concentrations and Lung Cancer Risk

To evaluate the association between directly measured blood protein levels and lung cancer risk, we used data on circulating concentrations of 85 proteins using the Olink platform in 732 lung cancer cases and 732 matched controls based on smoking status that were available within a parallel lung cancer early detection project (U19 CA203654). Conditional logistic regression models were used to estimate the association of proteins with lung cancer risk adjusting for age, body-mass index, and additional smoking characteristics (cigarettes per day and years smoked) and stratified by histology (Supplementary Methods). Test for trend of protein association with risk was across median concentration by quartiles.

#### Protein Tumor Expression and All-Cause Mortality Risk

The association of tumor gene expression in lung cancer patients with all-cause mortality in TCGA was estimated using Cox-regression to time from diagnosis to death as an underlying time variable. Models were additionally adjusted for age at diagnosis and stratified by sex, smoking status, and tumor-stage. Statistical analyses were performed using R (*TwoSampleMR*^30^, *Coloc*^25^; *hyprcoloc*^26^, The R project^31^).

## Results

### Identification of promising lung cancer proteins

#### Mendelian randomization Risk Analyses

In the initial MR analyses of 85 proteins for lung cancer risk, at an FDR threshold of 5%, we discovered an association with lung cancer risk for interleukin 18 (IL-18, N_SNP_=6 [5 *trans* and 1 *cis*], variance explained:4.3%, Figure 2). This association remained after pleiotropy-robust analyses: weighted median and MR-PRESSO and was directionally concordant using MR-Egger regression (eTable2). As specified *a priori*, IL-18 was therefore the only protein further investigated. A genetically predicted increment in blood IL-18 concentration (SD units) was associated with a reduced risk of lung cancer (OR_IVW_:0.85, 95%CI:0.79-0.92, N_SNP_=6; eTable1). The association between IL-18 and lung cancer risk appeared more prominent for patients diagnosed with adenocarcinoma (OR_IVW_:0.80, 95%CI:0. 72-0.89, N_SNP_ = 6; eTable2, Figure 3). No association was observed with other histological subtypes (eTable2). Additional sensitivity analyses revealed no evidence of horizontal pleiotropy. MR-Egger (eTable 3), Cochran’s or Rücker’s Q heterogeneity statistics (eTable 4) and openGWAS results (eTable 5) are shown in Supplementary Materials. Bidirectional MR analysis did not indicate an influence of lung cancer on circulating IL-18 levels (eTable 6). The only pleiotropic association between the IL-18 *cis*-SNP, rs5744249, was seen with heel bone mineral density (eTable 5). However, there was a low probability for a colocalized signal for IL-18 and heel bone mineral density at a locus around rs5744249 (7.8^-35^%) implying this may be due to confounding by LD. For *trans* IL-18 SNPs, proteome-wide association analyses pleiotropy was observed only for rs385076 and rs17229943 with Heme-Oxygenase 1 (results not shown); main results were materially unaltered when these SNPs were excluded (OR_IVW_:0.84, 95%CI:0.76-0.93, N_SNP_ = 4). Notably, no significant pleiotropy for IL-18 instruments was observed for its well-documented inhibitory protein, IL18-BP^32^, which may imply the IL18 MR risk estimates are not influenced by genetically elevated IL18-BP.

**Figure 2.**
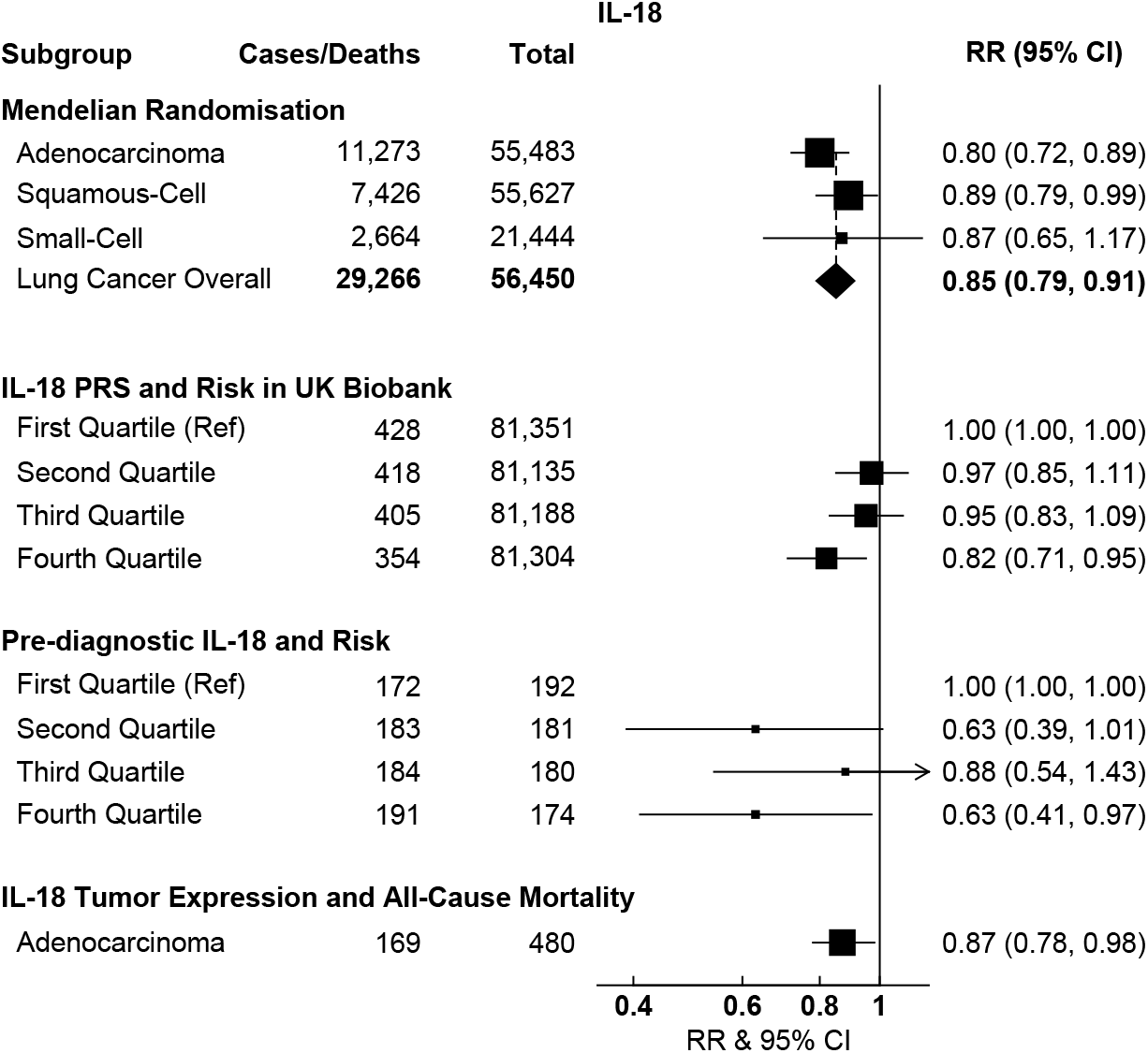
Volcano plot for the association of 85 inflammation and immune-related proteins with risk for lung cancer overall and by histological subtype. P values and odds ratios are based on Wald estimates for proteins with one SNP instrument or the inverse-variance weighted approach where multiple SNP instruments were available.

**Figure 3.**
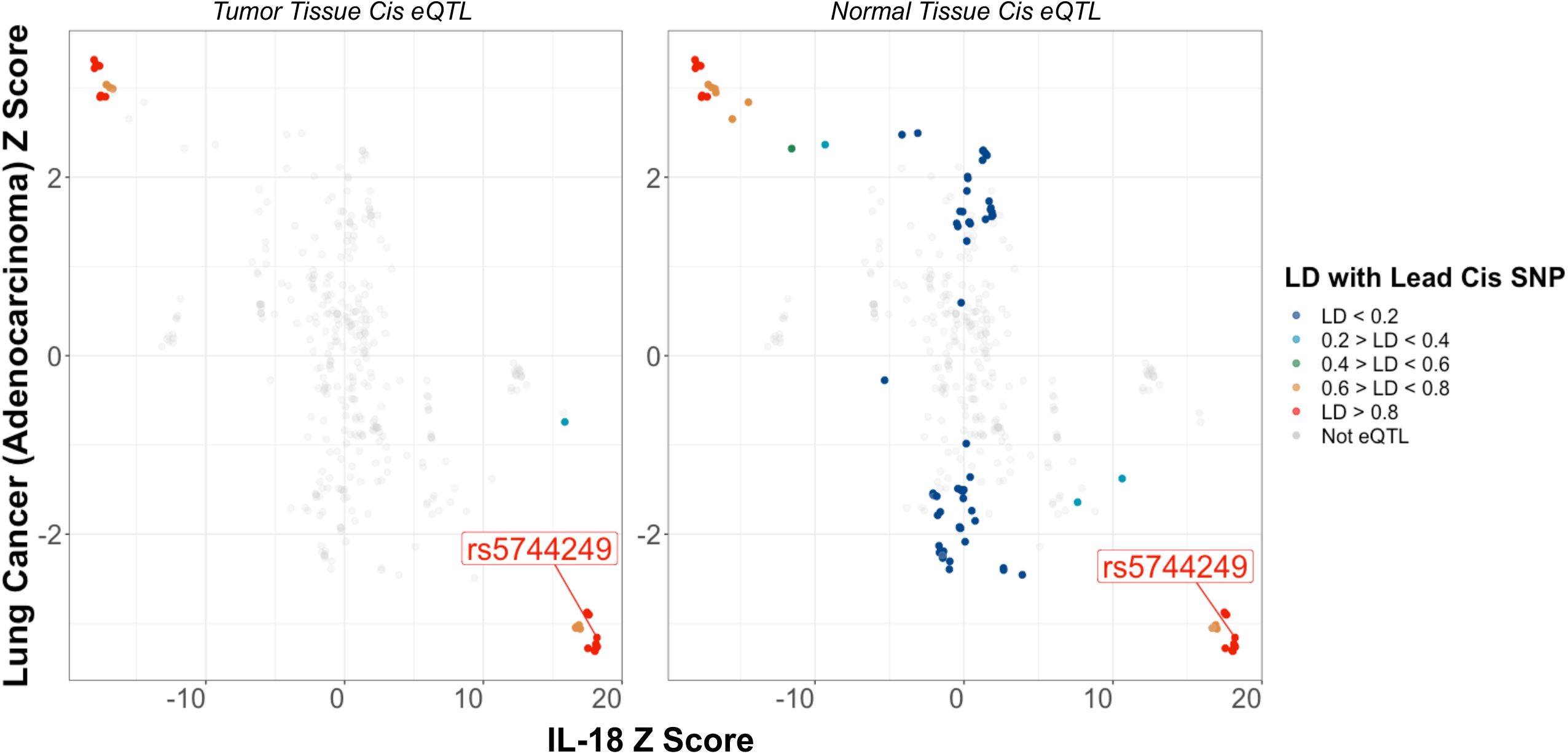
Summary of risk analyses for the association of IL-18 with risk for lung cancer from MR and all triangulation analyses: IL-18 PRS (by quartile) in UK Biobank using Cox-regression, pre-diagnostic IL-18 blood concentrations (by quartile with interaction for IL-18BP concentrations) using conditional logistic regression in a prospective nested case-control study, and IL-18 tumor expression (per SD) using Cox-regression in adenocarcinoma patients in the Cancer Genome Atlas.

The *cis*-pQTL for IL-18 is located upstream of the second exon adjacent to a promoter region. MR analyses using only this *cis*-pQTL supported our primary IL-18 MR results overall (OR_Wald_:0.87, 95%CI:0.79-0.94, N_SNP_ = 1; eTable2), and suggested a stronger association in adenocarcinoma (OR_Wald_:0.79, 95%CI:0.69-0.91, N_SNP_ = 1; eTable2).

#### Colocalization analyses

Little evidence was found to support colocalization for lung cancer overall (11%), small cell (1.7%), or squamous cell carcinoma (1.3%). However, there was moderate support for colocalization between IL-18 protein levels and risk for adenocarcinoma (colocalization probability 64%). To investigate further whether an association of IL-18 with adenocarcinoma acts via expression of IL-18 in lung tissue we additionally performed colocalization for IL-18 cis pQTL with IL-18 cis eQTL in normal lung tissue (99.8%); for IL-18 cis eQTL with adenocarcinoma risk (61.2%); and finally, a multivariable colocalization for cis pQTL and eQTL, and adenocarcinoma risk (89.8%, Figure 4 & Supplementary Figure 1). Although full summary statistics were unavailable for IL-18 tumor eQTL, there was considerable overlap for tumor and normal tissue significant eQTL in the lung. Together these results imply a shared genetic signal for IL-18 protein concentrations and IL-18 lung tissue expression with adenocarcinoma risk.

**Figure 4.**
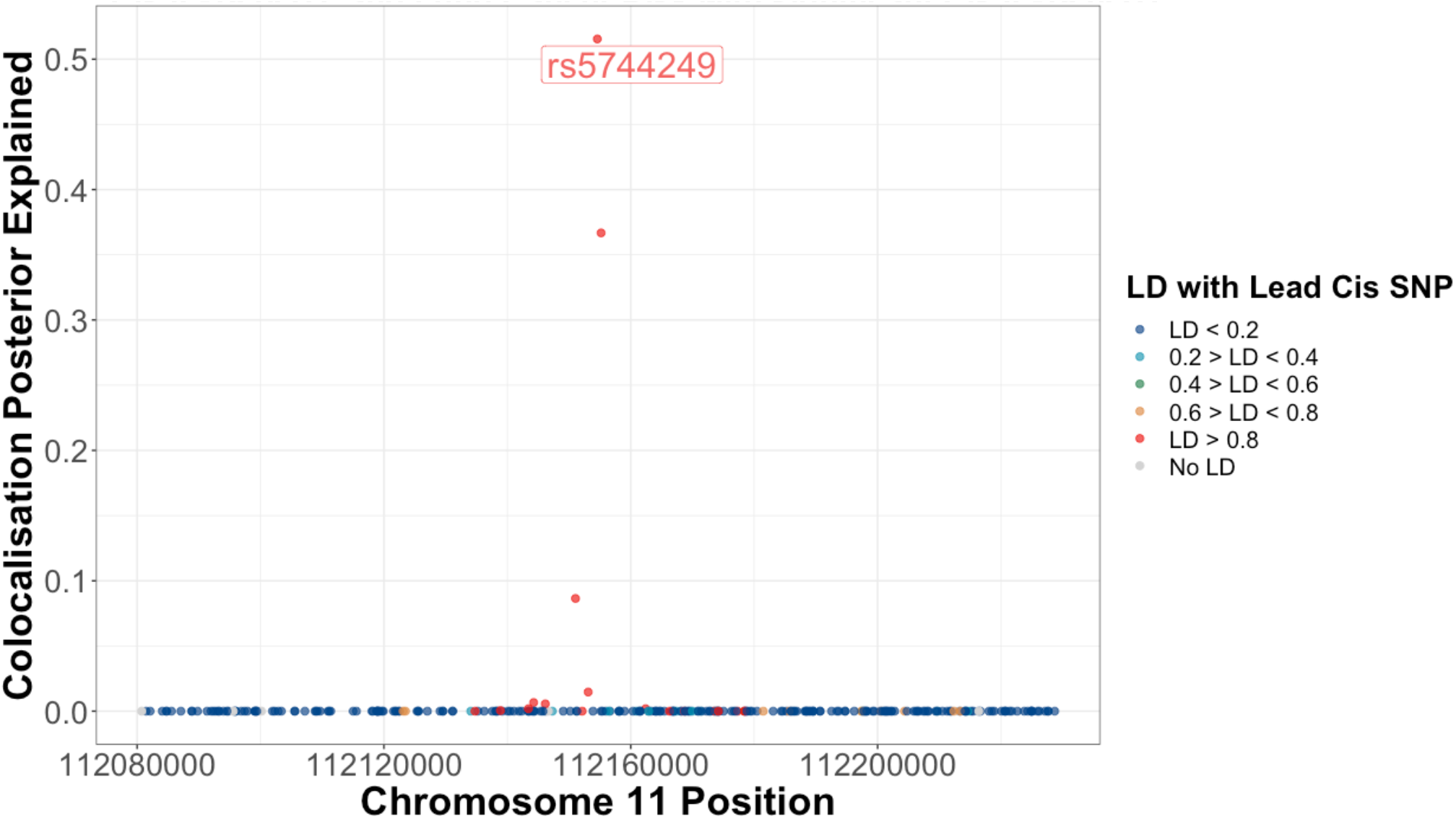
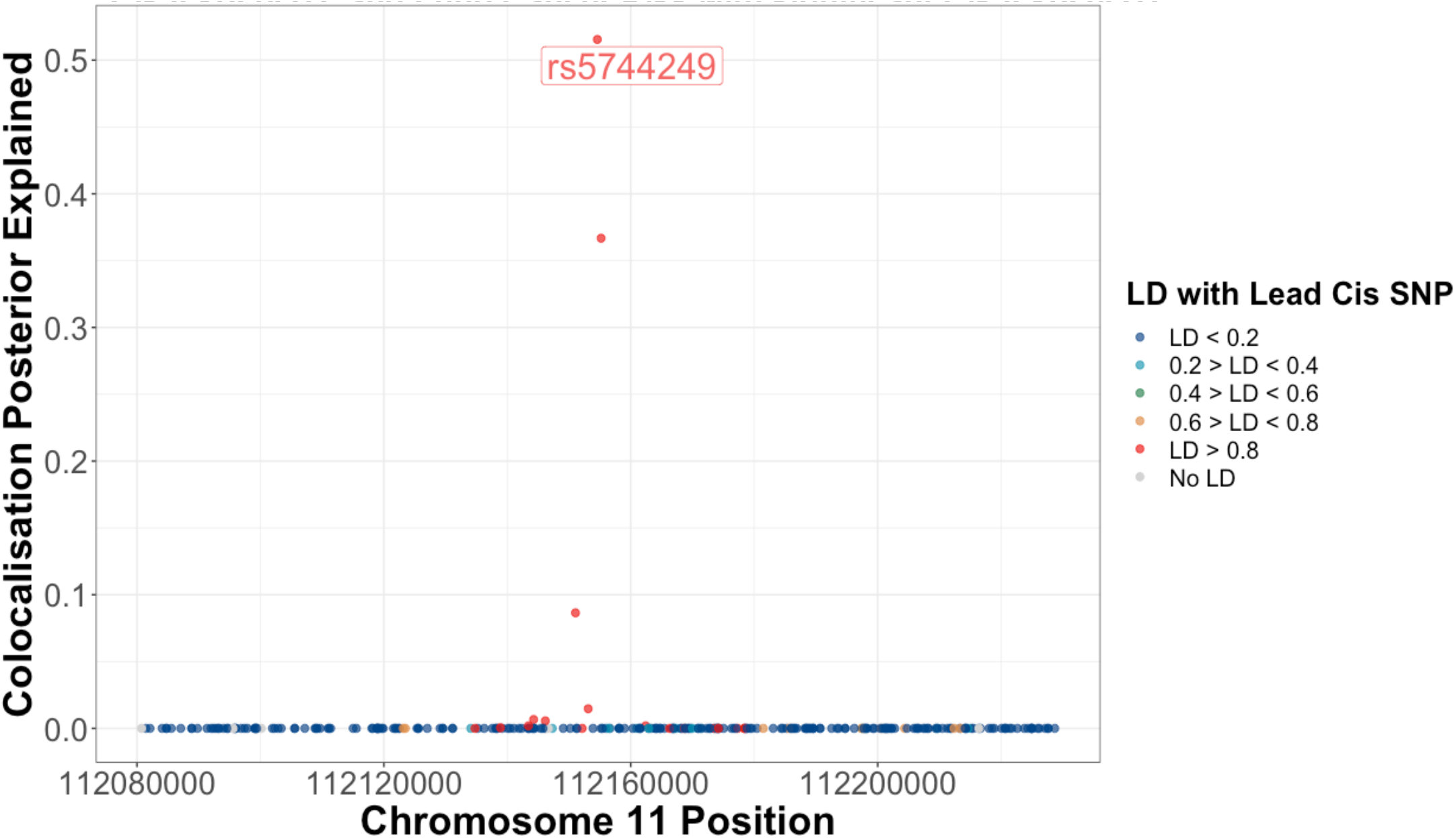
Colocalization plot for the association of SNPs with IL-18 levels and risk for lung adenocarcinoma within a 75kb region up and downstream of the IL-18 *cis*-pQTL (rs5744249). Functional annotations of IL-18 *cis*-eQTL SNPs in healthy and adenocarcinoma tumor tissue are colored as a function of their linkage disequilibrium with rs5744249. These plots highlight the consistent association of SNPs that are associated with lower IL-18 levels at this locus with higher risk for lung adenocarcinoma, which is more prominent among significant IL-18 *cis*-eQTL.

#### Triangulating evidence for promising lung cancer proteins Effect of IL-18 on the Tumor Microenvironment

From amongst immune traits available in TCGA, IL-18 levels have well-documented associations with Th1 cells^33,34^, CD8 T cells^32^, NK cells^32^, macrophages^35^, and Interferon Gamma Response^36^. We observed MR associations of genetically predicted IL-18 levels using the six pQTL instruments used in main risk analyses with an increase in pan-cancer tumor cells, including Th1 cells (p=0.004), CD8 T cells (p=0.01), NK cells (p=0.026), and macrophages (p=0.003). No clear association was observed for pan-cancer interferon gamma response (eTable6). Significant *cis-* pQTL (rs5744249) associations were observed for IL-18 with pan-cancer Th1 cells (p=0.03) and macrophages (p=0.005, eTable7). However, no strong evidence was observed in favor of colocalization between IL-18 levels and these characteristics (Th1 cell:34.4%, macrophages:10.2%).

Higher IL-18 gene expression in tumor tissue samples from 480 adenocarcinoma patients in TCGA was associated with a significantly greater tumor tissue CD8 T cell (beta per SD:0.11 [95%CI:0.02-0.21], p=0.02) and NK cell count (beta per SD:0.13 [95%CI:0.03-0.23], p=0.01), and IFN-γ pathway activation (beta per SD:0.24 [95%CI:0.15-0.34], p<0.001) after adjustment for age at diagnosis, gender, smoking status, and tumor stage. No significant association for IL-18 was observed with Th1 cell count (beta per SD:0.04 [95%CI:-0.06-0.14], p=0.4).

#### IL-18 PRS and Risk in UK Biobank

Polygenic predicted IL-18 (LDPred Score, variance explained:24%) was available for 326,583 UK Biobank participants that had no overlap with either the IL-18 protein or lung cancer GWAS, of whom 1,605 had been diagnosed with incident lung cancer. For cases, the mean follow-up time from baseline to diagnosis was 4.1 years (IQR:0.03-5.86 years)(eTable10). Cox regression identified an inverse association for higher genetically predicted IL-18 and risk for lung cancer (Hazards Ratio [HR] highest vs. lowest fourth:0.83 [95%CI:0.72-0.95], P_trend_=0.009; HR per SD:0.95 [95%CI:0.90-0.99], eTable13).

#### Pre-diagnostic IL-18 and Risk

Data from 732 cases and 732 matched controls identified within six prospective cohorts were included in the analysis. For cases, the mean time between pre-diagnostic blood collection and diagnosis was 1.6 years (IQR:0.8-2.4 years) (eTable11).

Base model conditional logistic regressions showed a positive association for direct pre-diagnostic blood measurements of IL-18 with lung cancer risk (OR highest vs. lowest fourth:1.39 [95%CI:1.02-1.90], P_trend_=0.04, eTable13). However, this association was attenuated after subsequent adjustment for smoking behaviors (cigarettes per day, years smoked, time since quitting) (OR highest vs. lowest fourth: 1.28 [95% CI: 0.92, 1.78], P_trend_ = 0.14, eTable13).

It is well known that IL18BP is a natural IL-18 inhibitor, affecting maturation and concentration of bioactive IL-18, which determines IL-18-stimulated immune responses via the IFN-γ pathway^32^. Therefore, we expect IL-18 to have a biological effect only when it is at a concentration greater than IL-18BP. This would ideally be modelled by a simple ratio, however, log2 relative quantification of proteins, as is available using the Olink platform, with differing natural abundancies do not allow for this ratio to be interpretable. Accordingly, we modeled the inhibitory effect of IL-18BP on IL-18 using an interaction term between IL-18BP and IL-18 with main effects retained in the model. This revealed an inverse association for IL-18 with lung cancer risk from smoking adjusted models in accordance with the MR analysis (OR highest vs. lowest fourth:0.63 [95%CI:0.41-0.96], P_trend_=0.03; OR per SD:0.91 [95%CI:0.83-0.98], eTable13, Figure3). No significant association was observed for IL-18 with lung cancer by histological subtype (eTable13).

##### IL-18 tumor expression and all-cause mortality in lung cancer patients

Similar to prospective analyses, there was no significant association for IL-18 expression with all-cause mortality after adjustment for age, gender, smoking status, and stage (HR per SD:1.05 [95%CI:0.91-1.21], p=0.5). Given that tumor expression data exist on a *raw* RNA count scale, a simple IL-18/IL-18BP ratio was employed to identify participants with likely bioactive tumor expression of IL-18. Similarly in the prospective risk analysis, Cox models found an inverse association between IL-18/IL-18BP with all-cause mortality among lung cancer patients (HR per SD: 0.87 [95% CI: 0.78-0.98], p=0.01, Figure 3).

## Discussion

We sought to identify novel inflammation-related proteins with a role in lung cancer aetiology by integrating results from large-scale GWAS, prospective cohorts, and tumor genomic analyses. The initial MR analysis queried 85 proteins and identified an inverse relation between IL-18 and lung cancer risk, and downstream analyses provided concordant evidence indicative of a protective role of IL-18 in lung cancer aetiology.

Specifically, we carried out an initial discovery analysis using MR to identify putative causal biomarkers, which revealed an estimated 15% reduction in lung cancer risk per standard deviation increase in IL-18 plasma levels. We also provide evidence that this association may act on risk locally via expression in lung tissue and potentially mediated by influencing the abundance of CD8 T cells and NK cells in the tumor microenvironment. The lung cancer susceptibility signal was validated in two prospective studies and one case-series using both genetically predicted and directly measured IL-18 levels. Polygenically predicted IL-18 in UK Biobank found a 17% lower risk for lung cancer among individuals with high IL-18. Similarly, direct IL-18 measurements in pre-diagnostic bloods were associated with a 37% reduced risk for lung cancer after accounting for the well-documented IL-18 inhibitory effects of IL-18BP. Lastly, we identified a 13% lower risk for all-cause mortality for higher IL-18 lung tumor tissue expression in adenocarcinoma patients after accounting for IL-18BP. These independent lines of evidence clearly indicate that IL-18 has an important role in mediating lung carcinogenesis.

IL-18 is a member of the IL-1 family, a group of 11 pro- and anti-inflammatory cytokines that are activated by NLRP3^37^, NLRP1^38^ and NLRC4 inflammasomes^39^. IL-18 drives MYD88 signaling^32^, which has been found to stimulate macrophages^40^, Th1 cells^33,34^, CD8 T cells and NK cells^32^, and drives interferon-gamma response^36^. We replicated the association of IL-18 with these immune characteristics using independent MR and tumor gene expression analyses finding evidence for an association for IL-18 expression with these immune cells local to lung tumor tissue. Colocalization of IL-18 pQTL with risk for lung cancer and lung tissue IL-18 expression was largely explained by the minor allele for the lead *cis*-pQTL/eQTL (rs5744249, 52%). Interestingly, we are believed to have introgressed this IL-18 elevating allele from Neandertals^41^, and so the protective effect of IL-18 may reflect immune response inherited from early homonid interbreeding^42^.

Therapeutically recombinant IL-18 (rIL-18) has a synergistic role with immune checkpoint inhibitors^43^ and may up-regulate immune pathways for tumorigenesis and progression^44^. However, even though rIL-18 is well-tolerated by patients in clinical trials^45^, it has been difficult to demonstrate the clinical utility of rIL-18. This has been attributed primarily to the negative regulatory role of IL-18BP in the bioactivity of circulating IL-18^46^. Indeed, a recent study that found IL-18 engineered as insensitive to IL-18BP has strong and consistent effects on T cell and NK cell activation in mouse tumors and acted to inhibit tumor progression^32^. It is therefore a strength of this study that no significant pleiotropy for IL-18 instruments was observed with IL-18BP implying our genetic MR discovery analyses was not affected by genetically higher IL-18BP. Further, based on the negative regulatory effect for IL-18BP on IL-18, it is in line with our current understanding of IL-18 function that an inverse association with lung cancer risk using direct measurements and all-cause mortality using tumor tissue expression was only observed after accounting for the inhibitory effect of IL-18BP.

Nonetheless, our study design was not without limitation. Firstly, pre-diagnostic blood measurements for IL-18 were among former and current smoking cases with a relatively short time between recruitment and diagnosis (max:3 years), which cannot protect against reverse-causation and precludes generalizing findings to never smokers. Further, the lack of treatment information available in TCGA did not allow us to explore the interplay between IL-18 tumor expression and survival by modern treatments known to act on potential IL-1 pathways, such as PD-L1 inhibitors. However, a re-analysis of CANTOS trial data found no association for the promising IL-1 beta antagonist Canakinumab for the reduction of IL-18^47^, which may imply IL-18 acts on risk for lung cancer via an alternative pathway to IL-1 beta.

In summary, we report multiple lines of evidence supporting a protective role of IL-18 in lung cancer aetiology. IL-18 may act to inhibit tumorigenesis via CD8 T and NK cell count local to lung tissue which is in line with the current understanding for IL-18 function. Altogether, these results highlight the potential for IL-18 as an aetiological biomarker for lung cancer and potential target for immune-oncology therapies.

## Data Availability

Summary statistics are freely available for protein GWAS from Folkersen et al. and for tumor immune characteristics from Sayaman et al.. Summary statistics for risk for lung cancer are available upon successful application to the ILCCO consortium. Individual-level patient characteristics for the Cancer Genome Atlas are available for download from dbGap. Individual-level protein data from the nested case-control studies are part of an ongoing study (INTEGRAL Project 2, U19 CA203654). These data will be available for analysis by investigators outside the study folllowing the date of publication of final results from the study's validation phase.

https://www.nature.com/articles/s42255-020-00287-2

https://www.sciencedirect.com/science/article/abs/pii/S1074761321000340

https://ilcco.iarc.fr

## Data Availability

Anonymized data will be shared by reasonable request from any qualified investigator.

---

**Supplementary Figure 1.**
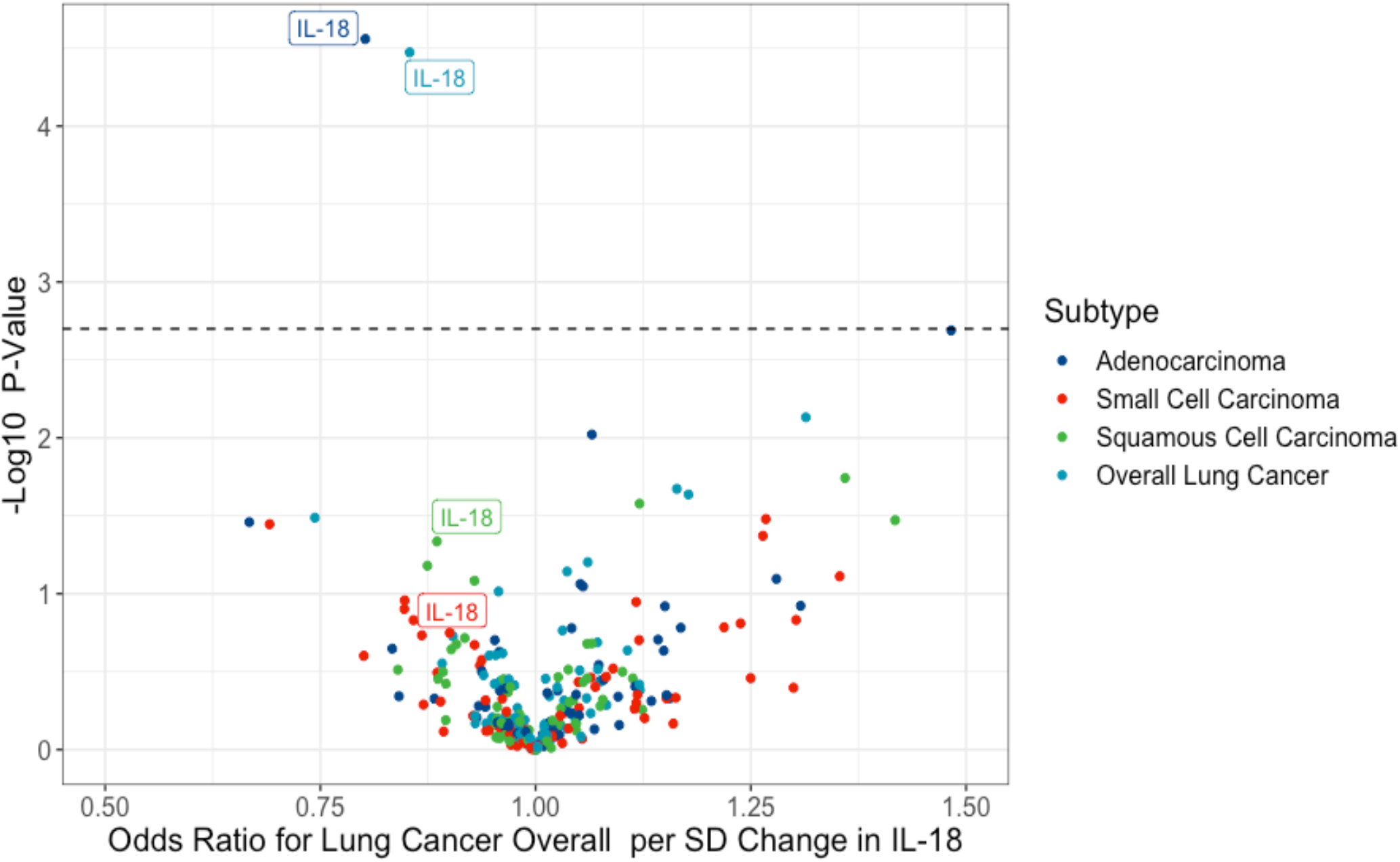
Proportion of the posterior believe for the colocalization of IL-18 protein levels, IL-18 healthy lung tissue expression, and risk for lung adenocarcinoma for each SNP in a 75kb window up and downstream of sentinel IL-18 *cis*-pQTL, rs5744249.

## Acknowledgements

This research was supported by funding from the US National Institutes of Health (INTEGRAL Program grant U19 CA203654) and Cancer Research UK (C18281/A29019). We would additionally like to acknowledge the SCALLOP and ILCCO consortia for generating the extensive summary statistics making the discovery analysis possible.

