## Supplementary Methods for "IL-18 and Lower Risk for Lung Cancer: Triangulated Evidence from Germline Predictions, Pre-Diagnostic Measurements, and Tumor Expression"

**Exposure and Outcome GWAS Description**

**-Inflammatory Protein GWAS**

This study consisted of up to 30,931 individuals from 14 studies with measurements on 90 proteins measured using the Cardiovascular-I panel on the Olink platform as part of the SCALLOP consortium (Average protein GWAS sample size: 17,747). GWAS analyses were conducted for inverse-rank normalised/standardised protein concentrations using an additive model in a linear regression with adjustment for population structure using principal components of ancestry in each SCALLOP study. All study results were combined using a fixed effects meta-analysis with inverse-variance weighting. Further details are available elsewhere^1^.

**-Lung cancer GWAS**

The study involved 85,716 participants (29,266 cases; 56,450 controls) from several genetic studies managed by the Transdisciplinary Research of Cancer in Lung of the International Lung Cancer Consortium (TRICL-ILCCO) and the Lung Cancer Cohort Consortium (LC3). GWAS analysis involved logistic regression models for each SNP (imputation was based on the 1000 Genomes Project Phase 3 panel) that included the 3 first principal components from a principal component analysis to infer genetic ancestry by genotype. Fixed effects meta-analysis with inverse-variance weighting was used to integrate results from the participating studies; description of all studies that took part of the meta-analysis, their inclusion/exclusion criteria, genotype quality control and detailed analytical plan are described elsewhere.^2^

**Protein-Tumor Immune Analyses**

**- Quality Control for Samples in the Cancer Genome Atlas**

The Cancer Genome Atlas^14^ is a joint effort between the National Cancer Institute and the National Human Genome Research Institute that characterised over 10,000 primary cancers across 33 cancer types. Patient samples were filtered to exclude the following TCGA^3^ non-rescinded annotations: "Item flagged DNU", "Administrative Compliance", "Item does/may not meet study protocol", "Qualified in error", "BCR Notification", "Normal tissue origin incorrect", "Subject withdrew consent", "Normal class but appears diseased", "Duplicate item", "Tumor tissue origin incorrect", "Tumor type incorrect", "Genotype mismatch", "Permanently missing item or object" and "Qualification metrics changed".

**Protein PRS and Lung Cancer Risk**

**-Study Samples and PRS method**

PRS analyses were conducted in the UK Biobank, a large prospective study started in 2006 with genotype information in over 500,000 participants age between 40 and 60 at recruitment. For this study, sample selection and SNP filters were non-white British ancestry, sex discrepancy, aneuploidy, and 2^nd^ degree relatives leaving 344,364 participants for analyses. Genetically predicted protein concentrations for proteins with significant MR associations were generated using the LDPred algorithm^4^ in Folkersen et al^1^ and estimated in UK Biobank using PRSice^5^.

**Pre-diagnostic Protein Concentrations and Lung Cancer Risk**

**-Study Samples**

Measurements for up to 1,182 proteins were available for a nested case-control study as part of an ongoing lung cancer protein project (U19 CA203654) using the Olink platform within six prospective cohorts. Pre-diagnostic measurements for proteins with promising MR results were extracted from these data to facilitate triangulation for lung cancer aetiology.

*EPIC*

The European Investigation into Cancer and Nutrition (EPIC) study is a large multi-centre prospective cohort that recruited participants between 1992 and 1998 and gather both questionnaire data and blood samples at recruitment from participants. For the case-control study, participants were taken from the 238,816 individuals from the centres in Netherlands, UK, France, Germany, Spain, and Italy who donated a blood sample at study recruitment. Further information regarding EPIC is provided elsewhere^6^.

*NSHDS*

The Northern Sweden Health and Disease Study (NSHDS)^7^ includes several prospective cohorts. The current study included study participants from the Västerbotten Intervention Project (VIP), which is a sub-cohort within NSHDS. In 1985, when VIP was started, all residents in the Västerbotten County were invited to participate by attending a health check-up at 40, 50 and 60 years of age. Participants completed a self-administered questionnaire and donated blood samples (fasting) for future research purposes.

*HUNT*

The Trøndelag Health Study (HUNT)^8^ is a large population-based cohort started in the 1980’s, covering 125,000 Norwegian participants from three sub-cohorts: HUNT1 (1984-86), HUNT2 (1995-97) and HUNT3 (2006-08). The study was primarily set up to address arterial hypertension, diabetes, screening of tuberculosis, and quality of life, however, this has expanded to include additional diseases.  The HUNT Study includes data from questionnaires, interviews, clinical measurements and biological samples (blood and urine).

*CPS-II*

The Cancer Prevention Study II (CPS-II)^9^ is a prospective mortality study of approximately 1.2 million American men and women recruited in all 50 states, the District of Columbia, and Puerto Rico beginning in 1982. Participants completed a questionnaire for lifestyle and anthropometric characteristics and 40,000 donated a blood sample at recruitment. During the 24 years of completed mortality follow-up currently available for this cohort (1982-2006), 491,188 deaths have occurred; cause of death has been obtained for 99.3% of all deaths.

*SCHS*

The Singapore Chinese Health Study (SCHS)^10^ is a population-based prospective cohort study of 63,000 Chinese men and women living in Singapore recruited between 1993 and 1998. The primary goal of this longitudinal cohort study is to elucidate the role of diet and its interaction with genetic and genomic factors in the causation of cancer and other chronic diseases. Each participant completed a lifestyle and anthropometry questionnaire and donated a blood and urine.

*MCCS*

The Melbourne Collaborative Cohort Study (MCCS)^11^ is a prospective population cohort of 41,513 residents of the Melbourne metropolitan area recruited between 1990 and 1994 with the aim of investigating the roles of diet and lifestyle in causing cancer and other non-communicable diseases. Participant completed a lifestyle and anthropometry questionnaire and a majority of participants also donated a blood sample.

- **Matching Criteria**

Controls were individually matched to the index case, cohort, study centre (where relevant), sex, date of blood collection (±1 month, relaxed to ±3 months for sets without available controls), and date of birth (±1 year, relaxed to ±3 years), and smoking status in four categories: former smokers with <10 or ≥10 years since quitting, and current smokers with <15 or ≥15 cigarettes smoked per day. Quartiles were fourths delimited in the controls.

- **Protein Measurements**

The abundance of proteins was measured in plasma or serum samples by multiplex proximity extension assay by Olink Proteomics AB (Uppsala, Sweden) using a Fluidigm Biomark reader (Fluidigm Corporation, USA). Paired case-control samples were randomly allocated over 96-well plates. Protein levels were measured as normalized protein expression (Ct values with corrections for assay variation) on the log_2_ scale. We excluded samples that did not pass laboratory quality control criteria (100% of values were missing, N = 3). As per advice, samples below the lower limit of detection (LOD) were replaced with LOD/sqrt(2)^12^. Additionally, we excluded samples that did not pass quality control criteria provided by the manufacturer.
